# Genetic evidence for the link of misophonia with psychiatric disorders and personality

**DOI:** 10.1101/2022.09.04.22279567

**Authors:** Dirk J.A. Smit, Melissa Bakker, Abdel Abdellaoui, Alex E. Hoetink, Nienke C.C. Vulink, Damiaan Denys

## Abstract

Patients with misophonia experience strong negative emotional responses to human-produced sounds at a level disrupting normal social interaction. The exact nature of the disorder remains a matter of debate. Here, we investigated the genetic aetiology of misophonia in order to understand contributing factors and shed light on the nosology of the disorder. For misophonia, we used an unpublished genome-wide association study (GWAS) from 23andMe on a self-report item probing a common misophonic symptom: the occurrence of rage when others produce eating sounds.. We used gene-based and to functional annotation analyses to establish its neurobiological determinants. Next, we calculated genetic correlations (r_G_) of this misophonia GWAS with a wide range of traits and disorders from audiology (tinnitus, hearing performance and hearing trauma), psychiatry, neurology, and personality traits. Misophonia was significantly correlated with tinnitus, major depression disorder, post-traumatic stress disorder, and generalized anxiety disorder (0.12 < r_G_ < 0.22). Stronger genetic correlations (0.21 < r_G_ < 0.42) were observed for two clusters of personality traits: a neuroticism/guilt and an irritability/sensitivity cluster. Our results showed no genetic correlation with ADHD, OCD, and psychotic disorders. A negative correlation with autism spectrum disorder (ASD) was found, which may be surprising given the previously reported comorbidities and the sensory sensitivity reported in ASD. Clustering algorithms showed that misophonia consistently clustered with MDD, generalized anxiety, PTSD and related personality traits. We conclude that — based on genetics — misophonia most strongly clusters with psychiatric disorders and specific personality profile that matches those for anxiety and PTSD.

## Introduction

Misophonia is a newly described condition, in which trigger sounds—such as chewing or breathing—provoke disproportionately strong and involuntary feelings of anger, anxiety, and/or disgust. When severe enough, these emotional responses (or the associated avoidance behaviour) may impede family relations and/or work life, resulting in patients seeking help from health care professionals. Diagnostic criteria for misophonia were provided in 2013 (Schröder et al., 2013) along with a clinical symptom screening instrument (the A-MISO-S). Recently, however, a consensus panel was unable to converge on a clear nosology for misophonia, and classify it as either a ‘psychiatric disorder’ or the more general ‘medical disorder’ (Swedo et al., 2022). The expert panel also concluded that knowledge on the genetic and neurobiological underpinnings of misophonia are lacking, and that further investigation is needed into the relation of misophonia with other disorders to better characterize misophonia. In addition, such research should also focus on the relation with psychological/personality traits as contributing factors.

Our present aim is to use genetics to fill in some of these gaps. This study is primarily based on the analysis of a Genome-Wide Association Study (GWAS) of a common misophonia symptom, namely, a self-report item on chewing-sound induced rage and analysed by 23andMe, Inc. Using this symptom as a proxy variable for misophonia, Fayzullina et al. (2015) reported one genetic locus that was significantly associated with the sensitivity to chewing sounds. This genetic locus, rs2937573, is intronic to TENM2 gene that plays a role in cell adhesion and is highly expressed in neurons, synapses, in various stages of brain development. However, a full functional annotation has not yet been performed. Our first aim is to perform this analysis, which may provide insights into the neurobiological underpinnings of misophonia.

Our second aim is to determine the association between the genetics of the misophonia symptom with the genetics of many other traits. It is known that genetic aetiology of psychiatric disorders (and many other disorders and traits) show pervasive correlations (Bulik-Sullivan et al., 2015). This overlap shows strong clustering (Lee et al., 2019) across psychiatric disorders, for example, a substantial degree of overlap (genetic correlation r_G_=0.31) was reported between obsessive compulsive disorder (OCD) and bipolar disorder (BIP). Moreover, shared genetics extends to substance use disorders (Abdellaoui et al., 2020) and non-psychiatric variables such as socio-economic status, which has important consequences for nosology and identification of contributing factors (Marees et al., 2020b). Inspecting genetic correlations and placing misophonia in a network of disorders and traits will aid its nosology.

We selected a list of 44 traits and disorders for our genetic correlation analysis. Based on the phenotypic comorbidities of misophonia with psychiatric disorders, it seems most likely that misophonia will show significant genetic correlations with major depression, ASD and ADHD (Jager et al., 2020), possibly also with OCD and Tourette’s syndrome (Webber et al., 2014; Webber and Storch, 2015). In addition, we expect misophonia to correlate with personality dimensions (Jager et al., 2020). Therefore, personality traits will be added to the list of GWAS that may classify misophonia. A second group of disorders and traits comes from the field of audiology. Initially defined in this field as a form of decreased sound tolerance (Jastreboff and Jastreboff, 2001, 2014), misophonia may bear relation to audiological disorders and related traits. Finally, we added several traits that putatively bear relation to misophonia. Neurological traits may reflect neuronal excitability; cortical measures of the limbic cortex (viz., mean insula surface area and thickness) (Grasby et al., 2020) were included based on the Autonomous Sensory Meridian Response (ASMR) hypothesis of misophonia (McGeoch and Rouw, 2020). Finally, educational attainment is known to correlate with many psychiatric disorder as well as audiological performance measures, and was added for this reason.

## Methods

### GWAS summary statistics

The source GWASs are studies from 23andMe, UK Biobank and the Psychiatric Genomics Consortium. Supplemental Table 1 shows an overview of all the disorders and trait GWASs used, their sample sizes, and their source, and specifics on the measurement. All psychiatric disorder GWAS are case-control GWAS with ascertained samples from the PGC.

A case-control GWAS for misophonia based on the proposed clinical criteria does not yet exist, nor was the condition assessed in the UK BioBank. However, Fayzullina et al. (Fayzullina et al., 2015) published a GWAS on the self-reported item of “Does the sound of other people chewing fill you with rage?” in 80607 subjects from the general population, including only subjects who answered yes or no to the question. The prevalence of a positive answer to this question was 22%, which is likely to be an overestimation of the clinical prevalence of misophonia, although no population prevalence numbers for misophonia are currently available.

Wherever possible, European ancestry versions of the summary statistics were selected, since results from other ancestries may bias the results. There was no selection on gender. A total of 43 traits were compared with misophonia in this study. These traits were categorized in Audiological (N=10, including tinnitus and hearing performance traits), Psychiatric (N=11) and Personality (N=15). The remaining 8 traits were added to the category “Other” and included various neurological disorders (Alzheimer’s, Parkinson’s, epilepsy), insula measures (surface area and thickness), and socioeconomic factors Educational Attainment and Townsend Index. Several UK Biobank trait GWAS (for personality traits) were obtained from the NealeLab GWAS collection web page (NealeLab, 2018).

### GWAS of hearing traits

For several audiological traits a GWAS using the UK Biobank data was performed. To establish whether hearing problems may play a role, a quantitative GWAS was performed on the hearing test results (field 20019 and 20021, “Speech perception threshold” left and right ear using the speech-in-noise test). Additional case-control GWASs were performed for Hearing aid, Hearing problems, and Loud music exposure (fields 3393, 2247, 4836).

For tinnitus (UK Biobank field 4803, “Do you get or have you had noises (such as ringing or buzzing) in your head in one or both ears that lasts for more than 5 minutes at the time?”), two analyses were performed, one for “Ever Tinnitus” (combining all values from “yes, but not now, but have in the past”). In addition, one GWAS was estimated for “Current Tinnitus” (combining all values from “some of the time” and up, with the “yes, but not now, but have in the past” values removed). The Current Tinnitus GWAS was repeated for subjects with good hearing (below −5.5 dB on the hearing test, see above) to test the genetics of tinnitus without functional hearing loss (Dawes et al., 2014).

### Genetic annotation

We used the FUMA web-application (Watanabe et al., 2017) to perform gene based analysis with MAGMA based on chromosomal position. In addition, we identified expression quantitative trait loci (eQTLs) for different tissues from the GTEx (Lonsdale et al., 2013), BRAINEAC (http://www.braineac.org), and eQTLgen (Võsa et al., 2021) resources. Table S1 gives an overview of the tissues selected for the analyses.

We subsequently ran a Transcriptome-Wide Association Study (TWAS) for the GTEx tissues (version 8) using the FUSION software (Gusev et al., 2016). The TWAS resulted in a test for each tissue by gene combination reflecting the genetic association of misophonia with a gene for that particular tissue. Significance levels of these tests were FDR corrected across all tested genes within a tissue. These were further corrected for the multiple tissues: To correct for multiple testing of expression profiles across tissues—which are expected to highly correlate—we estimated the independent degrees of freedom of the cross-tissue imputed expression correlation matrix with spectral decomposition (Nyholt, 2004; Li and Ji, 2005). This number was used as a Bonferroni correction factor. The correlation matrix for input into the spectral decomposition was based on pairwise complete TWAS z-scores.

### Genetic correlations

To calculate the genetic correlation between the set of 44 GWASs, the package GenomicSEM (Genomic structural equation modelling, Grötzinger et al., 2019) was used in R (version 4.0.3, The R Foundation for Statistical Computing, 2020). With this package, pairwise bivariate LD score regression analyses were performed using the recommended settings across all traits (Bulik-Sullivan et al., 2015). SNP filtering settings were HWE p>1E-8, MAF>.01 insofar available, and were downsampled to HapMap 3 excluding the MHC region for the subsequent genetic correlation calculation (Bulik-Sullivan et al., 2015).

For visualization of the genetic correlations R-package corrplot (version 0.86) was used, using hierarchical clustering to order the traits. Before clustering, we identified traits that were reverse coded—that is, all traits that reflect positive aspects such as friend satisfaction were reversed. This was done by entering the full genetic correlation matrix into an eigen decomposition and extracting the loadings on the first unrotated principal component. Of traits with substantial negative loadings (below -.05) all genetic correlations were negated. All p-values were corrected for multiple testing using the False Discovery Rate control (Benjamini and Hochberg, 1988).

### Graph clustering

R package iGraph (version 1.2.6, Csardi & Nepusz, 2006) was used to determine the clustering of all traits included in the analysis using the Louvain method (Blondel et al, 2008). The method optimizes the modularity index q, which indexes the relative size of within cluster strength of the within-cluster strengths compared to the between-cluster strength, where strength was defined as the genetic correlation between trait pairs.

To establish the consistency of clustering we used the data provided by GenomicSEM to resample the genetic correlation matrix. A fuller account of this procedure is provided in the supplemental methods. In short, GenomicSEM provides variability (standard errors) of the SNP-h2 and SNP-covariance estimates (990 in total for the 44 traits) together with the covariation between estimates. This matrix (a 990 × 990 matrix for 44 traits) was used as the ‘sigma’ matrix in the R package mvrnorm, with the estimates (genomicSEM matrix S) themselves as the ‘mu’ parameter. This provided a set of 1000 samples with the resampled estimates in a single row with the correct mean value, variability, and covariability between the resampled estimates. Each resampling was reordered into a genetic correlation matrix, and reassessed with the Louvain clustering method. Finally, we counted the number of times pairs of traits were grouped in the same cluster to assess the consistency of clustering.

## Results

### Misophonia GWAS and annotation

Figure 1 shows the Manhattan plot for the misophonia GWAS (see also Fayzullina et al.). The top SNP rs2937573 was highly significant (p= 2.58 × 10^−43^) is near the TENM2 gene (intergenic in build37; intronic in build38). None of the SNPs within the LD block (cutoff r^2^ = 0.6) is an known eQTL for a gene, but several deleterious SNPs are present in the region, of which rs2915860, rs7728595, and rs2915858 were present in the main GWAS and significantly associated with misophonia (p<=1.28 × 10^−11^; CADD=>15.36). Remaining SNPs in the LD block with CADD>12.37 are listed in supplemental table 2 (Kircher et al., 2014). Opentargets.org and GWAS catalog lookup of the top SNP and SNPs in LD reported a link with adolescent scoliosis (Liu et al., 2018) and ‘Time spent watching TV’ (UK Biobank item 1080, analysis Neale v2).

**Figure 1.**
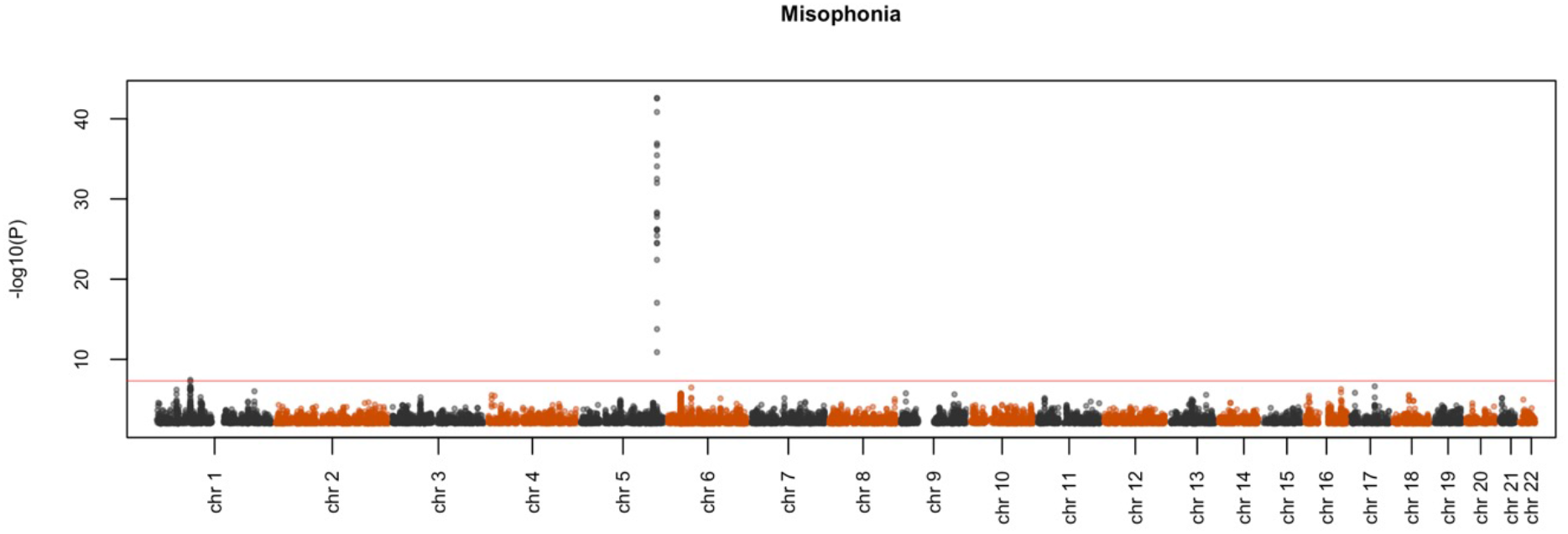
Manhattan plot from the Fayzullina et al. white paper from 23andMe. Results are 697384 associations from genotyped SNPs of N=80607 participants who reported either yes (N=17606) or no (N=63001) to the question: “Does the sound of other people chewing fill you with rage?”.

A second independent hit (rs7522520, p= 3.57 × 10^−8^) was located on chromosome 1 near pseudogene RN7SK. Supplemental table 2 shows the SNPs in the LD block of rs7522520 (cutoff r^2^ = 0.6) as reported in GWAS catalog (MacArthur et al., 2017), which includes a variety of traits in the wellbeing spectrum. One SNP within the LD block is an eQTL for NEGR1 (*Neuronal growth regulator 1*; rs6656687, eQTLgen blood tissue cis-eQTL, p=1.94 × 10^−23^). This SNP was not tested in the original GWAS. The LD block covered 22 SNPs with CADD score>12.37; none were significantly associated with misophonia.

FUMA positional gene-based analysis with MAGMA showed one Bonferroni corrected significant gene: TMEM256 (corrected p=0.0257) on chromosome 17.

### Expression analysis

We performed a Transcriptome-Wide Association Study analysis (TWAS) using 10 GTEx brain tissue and 1 whole-blood expression profiles. To correct for multiple testing, we applied FDR to adjust for testing across many genes. Across tissues, we further applied Bonferroni correction to the significance threshold. Since MatSpD (Nyholt, 2004) estimated df=3.84 as the effective degrees of freedom of the TWAS effects across tissues, alpha=0.0130 was used as the significance cut-off value. The TWAS revealed that *TFB1M* expression in Hippocampal tissue was the only significant effect (FDR-p=0.0055). *TFB1M* is located on chromosome 6 and encodes for one of several proteins that regulate mtDNA transcription and replication, and is associated with mitochondrial nonsyndromic sensorineural deafness, and drug-induced hearing loss (Bykhovskaya et al., 2004; O’Sullivan et al., 2017). *ECE2* gene expression in Putamen was close to significance (FDR-p=0.0168, *n*.*s*.).

### Misophonia genetic correlations

Misophonia showed substantial SNP heritability (h^2^=8.5%, z=10.5, p= 9.2 × 10^−26^). The LD-score regression intercept was 1.005 (SE=0.0088; p>0.05), indicating excellent control of inflation due to stratification. The genetic correlations of misophonia with the selected traits are shown in figure 2, clustered by category and ordered by magnitude. The full heatmap of the correlations is provided in figure 3.

**Figure 2.**
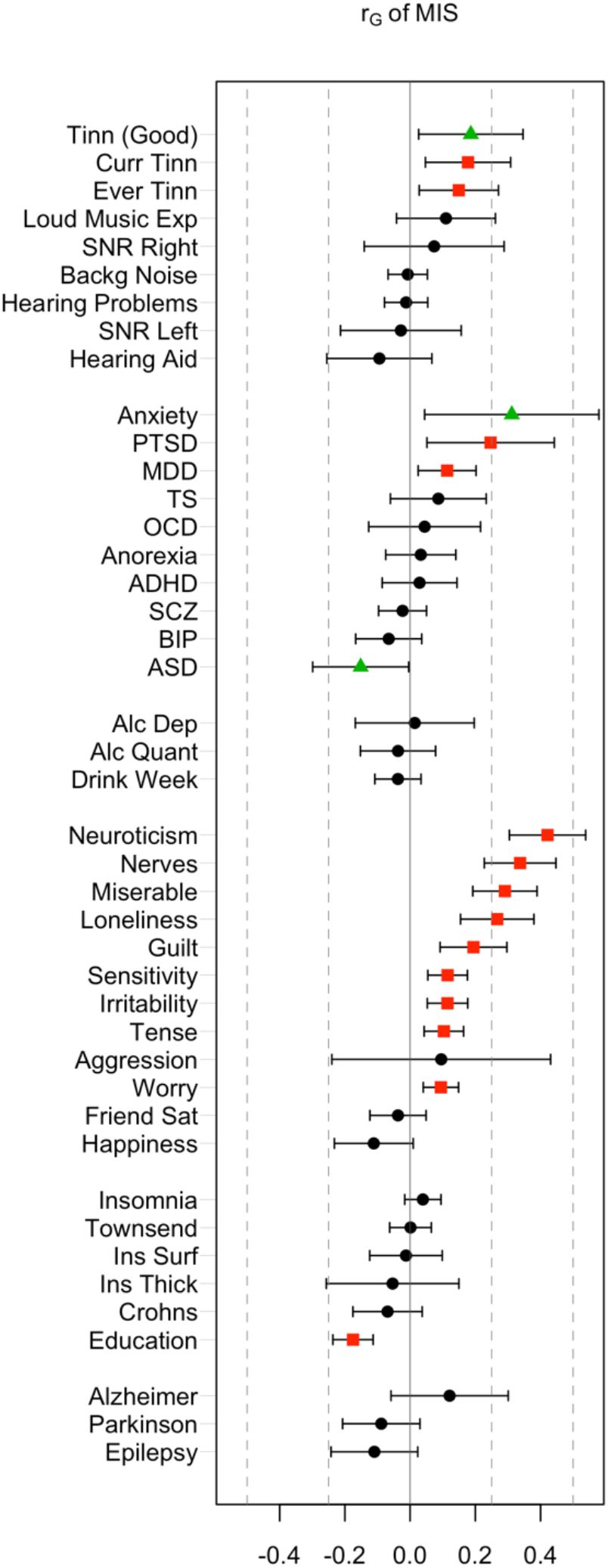
Genetic correlations of misophonia symptoms with a range of behavioural traits and disorders. FDR-corrected significant effects (red) were observed in most categories (audiological, psychiatric, personality, and miscellaneous traits). Nominal significance (green triangles) was observed for Anxiety, ASD, and ‘Tinnitus in good hearing’. FDR adjusted significant correlations were observed for Current Tinnitus, Ever Tinnitus, PTSD, MDD, a range of internalizing and externalizing traits, and educational attainment (red).

**Figure 3.**
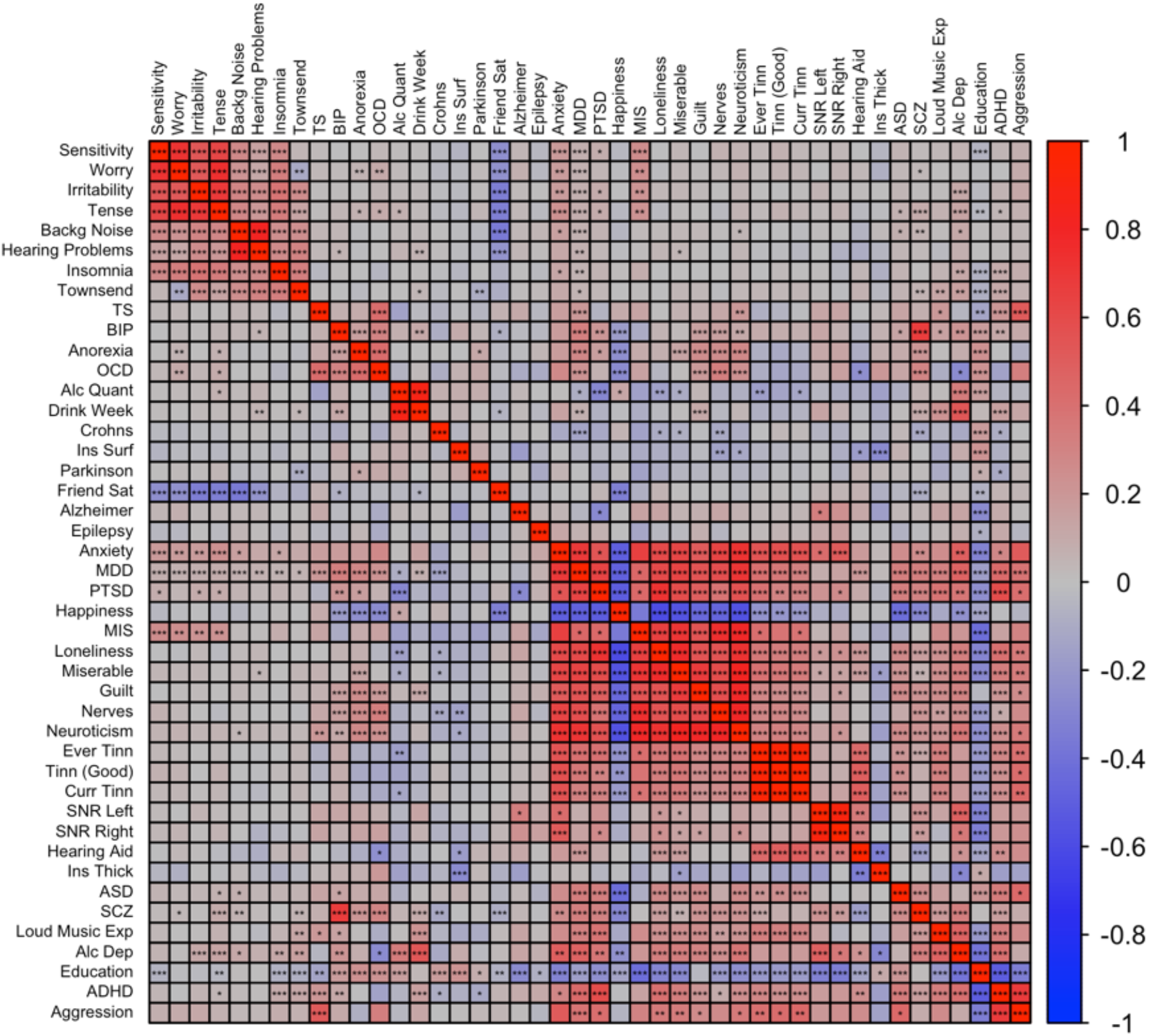
The full genetic correlation heatmap ordered using hierarchical clustering. Significance was based on FDR-corrected p-values across the full lower diagonal of the matrix. *p<0.05, **p<0.01, ***p<0.001, FDR adjusted across all traits

#### Misophonia to audiology correlational pattern

Misophonia showed moderate but significant positive correlations in the range from 0.15 to 0.19 with tinnitus (specifically, ‘Ever tinnitus’ and ‘Current tinnitus’). Correlations with other audiology traits were not significant. In contrast with misophonia, the three Tinnitus traits showed clear overlap with hearing trauma variables (Current Tinnitus: r_G_=0.51 for ‘Loud music exposure’; r_G_=0.29 for ‘Hearing Aid’). This is consistent with the early observations that misophonia is unrelated to hearing performance and/or hearing loss, whereas tinnitus often arises after hearing trauma (Langguth et al., 2013; Moore et al., 2017). ‘Loud Music Exposure’ is even likely to have a causal effect on tinnitus (Moore et al., 2017). Like misophonia, tinnitus was unrelated to hearing performance (SNR left or right), which seems at odds with tinnitus’ correlation with ‘hearing loss’, but may be due to the fact that most of the SNR trait variation within normal hearing range is unrelated to hearing problems (figure 4). This is visible as moderate correlations between SNR left and right with ‘hearing aid’ (r_G_=0.37 and r_G_=0.34 for left and right SNR respectively), and nonsignificant correlations with ‘hearing problems’.

**Figure 4.**
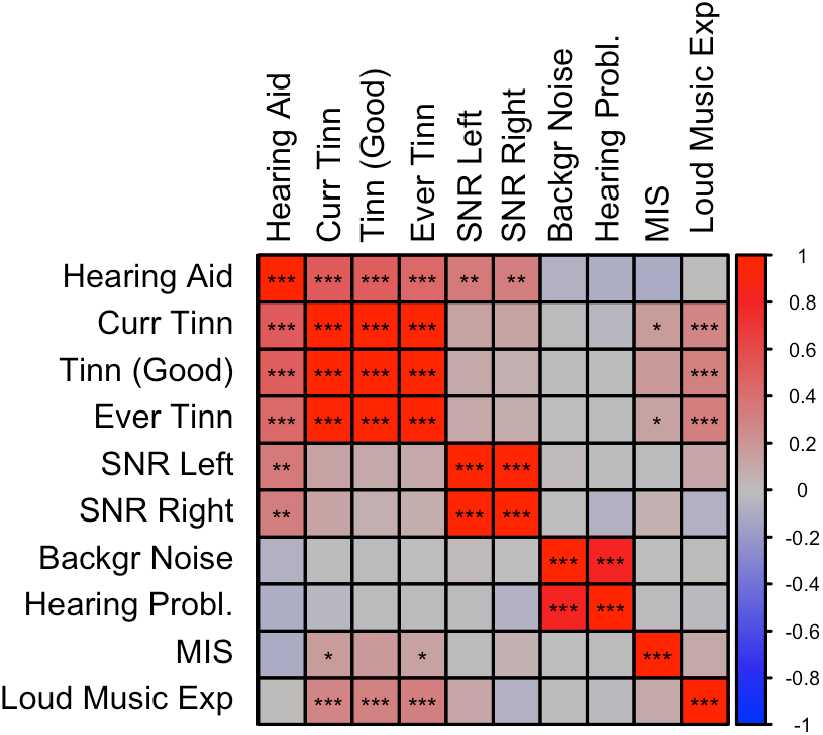
Correlation Plot between Misophonia and Audiological traits. SNP-based genetic correlations were calculated between Misophonia and 10 audiology traits using LDSC. *= p<0.05, **= p<0.01, ***= p<0.001, FDR adjusted across all traits combinations. See Supplementary Table 1 for abbreviations.

Interestingly, ‘Loud Music Exposure’ showed a pattern of correlations that was very similar to that of misophonia (i.e., moderate correlations with tinnitus variables and low correlations with the remaining traits), however, the <u>direct</u> correlation between ‘Loud music Exposure’ and misophonia was low and not significant. This further suggests that misophonia is not related to hearing problems, but reflects shared etiology with tinnitus via other (psychological) traits (Pattyn et al., 2016).

#### Misophonia and psychiatric liabilities

Consistent with the observed phenotypic correlations (i.e., comorbidities), we observed significant positive correlations between misophonia and MDD (r_G_=0.11, uncorrected p=0.012, FDR-p= 0.045) and PTSD (r_G_=0.25, uncorrected p=0.013, FDR-p=0.045). The strongest correlation was with Anxiety (r_g_ = 0.31, uncorrected p=0.022). Surprisingly, there was a (nominally) significant <u>negative</u> correlation between ASD and misophonia (r_G_=-0.15, uncorrected p=0.044, FDR-p>0.05).

#### Misophonia and psychological/personality traits

Misophonia was significantly positively correlated with Guilt, Loneliness, Miserableness, Nerves, Neuroticism, Irritability, Sensitivity, Tense feelings, and Worry. The strongest correlation was with Neuroticism (r_g_ = 0.42, uncorrected p=2.0×10^−12^, FDR-p=7.5×10^−11^). There were no negative correlations. A positive correlation with aggression did not reach significance, possibly due to the wide confidence intervals.

Figure 5 shows a correlation plot for a selection of above traits (i.e. significantly correlated to misophonia) plus the MDD, PTSD, and Anxiety liability traits that highly correlate with the selected psychological traits. From the correlational pattern two clusters appear, where Guilt, Nerves, Loneliness, Miserable, Neuroticism correlate highly. Irritability, Sensitivity, Tense and Worry formed a second cluster. Psychiatric disorders (generalized anxiety, PTSD, and MDD) clustered with the neuroticism/guilt cluster, but also showed significant overlap with the irritability/sensitivity cluster. Misophonia closely followed the pattern of correlations of the psychiatric disorders, as it clusters in the neuroticism/guilt cluster but also shows moderate genetic correlations with the irritability/sensitivity cluster.

**Figure 5.**
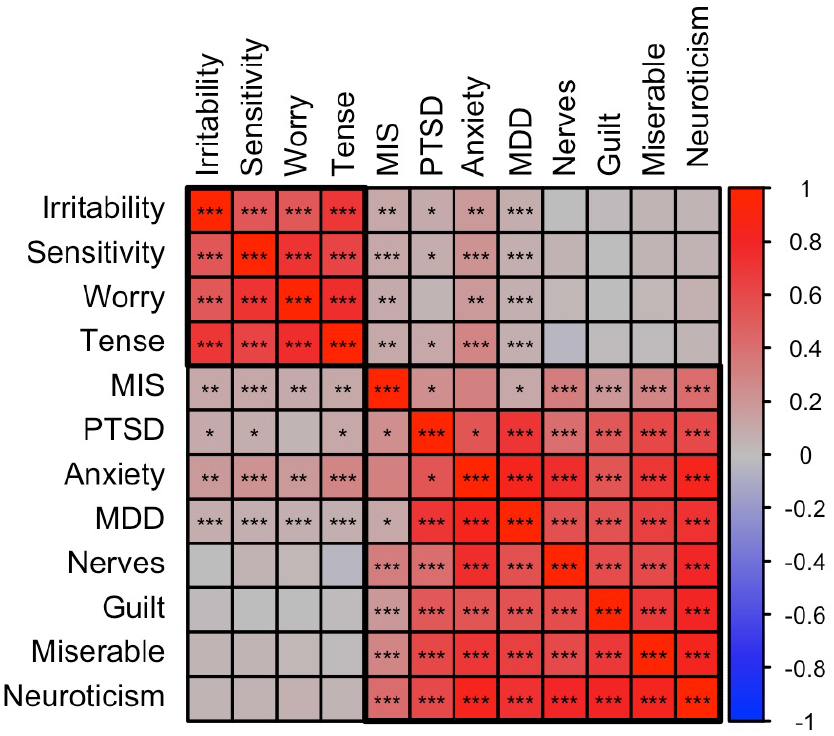
Correlation Plot between Misophonia, Psychiatric, and Psychological traits. SNP-based genetic correlations were calculated between Misophonia, Tinnitus (Current), and 12 selected traits shows the membership of two main clusters identified using hierarchical clustering, putatively called the irritability and neuroticism clusters. The neuroticism cluster holds most internalizing traits. The irritability cluster also holds sensitivity, tenseness, and worry. The psychiatric traits are clustered with the neuroticism cluster but all showed significant positive correlations with the irritability cluster. Misophonia closely followed the psychiatric traits. Tinnitus showed a pattern different from misophonia with high correlations with the neuroticism cluster but no significant correlations with the irritability cluster. *= p<0.05, **= p<0.01, ***= p<0.001, FDR adjusted across all traits combinations. See Supplementary Table 1 for abbreviations.

#### Misophonia and remaining disorders and traits

Environmental variables of social and living conditions as well as important cognitive traits are genetically correlated to some psychiatric disorders, including substance use disorders (Abdellaoui et al., 2019, 2020; Marees et al., 2020a). We therefore investigated ‘Townsend Index’—an index of impoverished living environment—and ‘Educational Attainment’ (EA) as possible contributing factors for misophonia. These variables are known to have a genetic component (Abdellaoui et al., 2019). Of these, misophonia showed a highly significant negative correlation with Educational Attainment (r_G_=-0.18, FDR-p=2.8*10^−8^). The Townsend index did not show a significant correlation (r_G_=0.00, p>0.05).

Educational attainment (EA) is known to have cognitive (IQ) and noncognitive factors (personality, environment). A recent article parsed genetic variance into these constituent parts, and reported that the genetic correlations between Educational attainment (EA) and psychiatry changed (Demange et al., 2021). Genetic correlations of EA with SCZ, Bipolar, Anorexia, and OCD were lower for the cognitive EA variance than for the noncognitive EA variance, even changing sign for SCZ and Bipolar. For misophonia, the reversed pattern was observed. Misophonia correlated significantly with noncognitive EA (r_G_=-0.162, SE=0.039, p=3.5 × 10^−5^) and a genetic correlation closer to zero was found for cognitive EA (r_G_=-0.071, SE=0.039, p=0.07).

There were no significant correlations of misophonia with the neurological disorders and insula measures.

### Misophonia falls in a personality/psychiatric cluster

The graph based on genetic correlations is shown in figure 6. Graph clustering showed that Misophonia clusters with psychiatric disorders anxiety, autism spectrum disorder, PTSD, major depression, anorexia nervosa, OCD, schizophrenia, and bipolar, and with psychological traits Guilt, Miserableness, Loneliness, Neuroticism, Nerves, and Happiness. Monte-Carlo resampling of the genetic correlation matrix and recalculating the clustering revealed that the membership of misophonia into this cluster was highly consistent, but this was not the case for all traits and disorders within that cluster. Misophonia clustered 95% of the samples with PTSD, MDD, Guilt, Happiness, Loneliness, Miserableness, Nerves, and Neuroticism. Cluster concordance was slightly less consistent with anxiety at 87% and with ASD at 61%. Note that the clustering with ASD is based on the consistently negative correlation with misophonia as sign is disregarded in the clustering algorithm. Anorexia, schizophrenia, and bipolar disorder clustered less than 41% of the time with misophonia. Figure 7 shows the full cluster concordance matrix.

**Figure 6.**
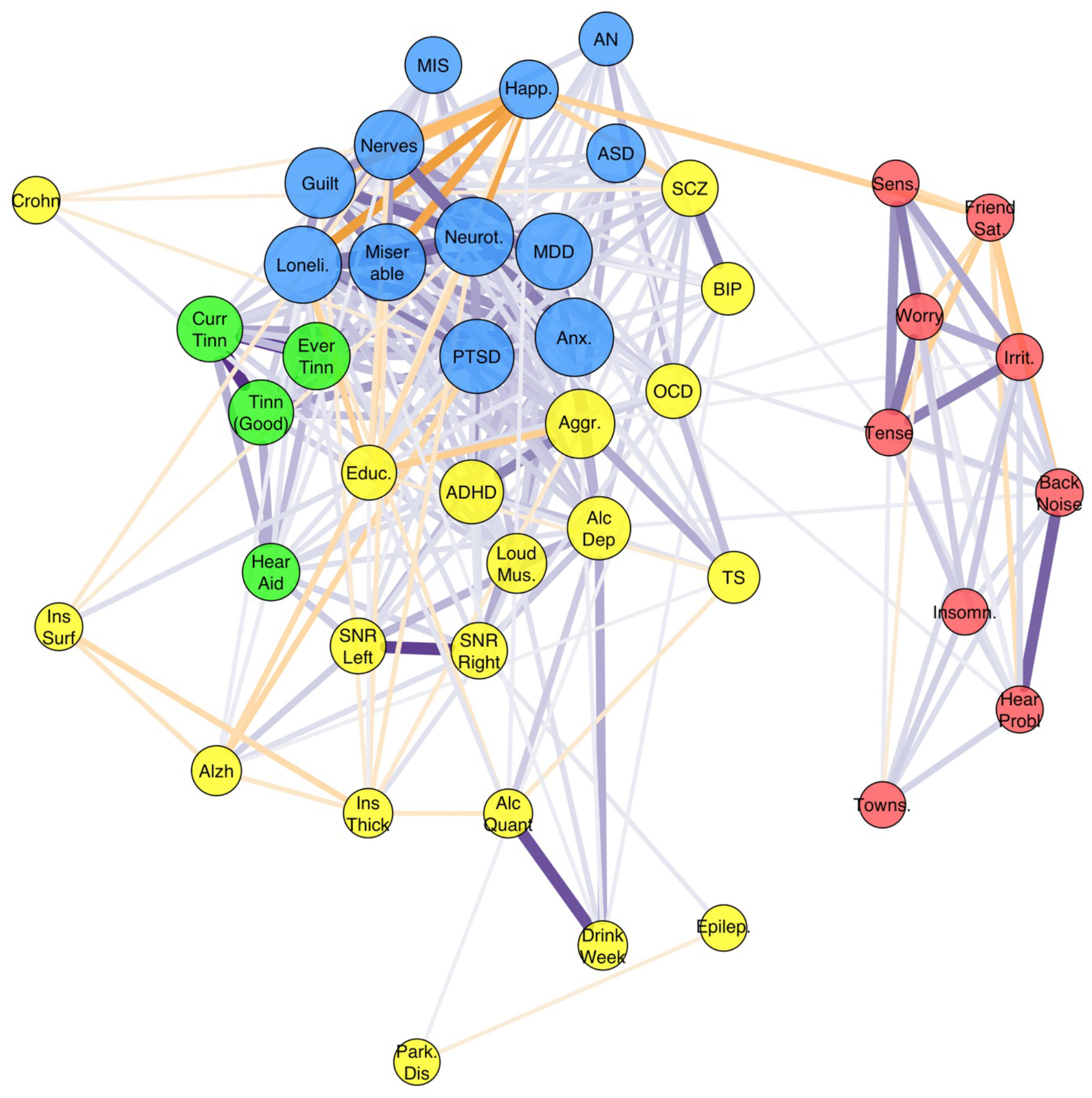
Graph of the full genetic correlation matrix. Vertex colours are based on Louvain clustering algorithm. Per vertex, only the top 10 edges are shown that are over 0.10. Node size is based on the eigencentrality of the trait calculated from the weighted correlation matrix (absolute values). Misophonia clustered with the Depressive disorders cluster (MDD, PTSD, and Anxiety) which also holds related personality traits (including neuroticism, guilt, miserableness, loneliness) (blue). Irritability and related traits (worry, sensitivity, tense) cluster with hearing problems (without or with background noise), insomnia, friendship satisfaction, and Townsend index (red). Tinnitus traits clustered with ‘hearing aid user’ (green). The remaining cluster (yellow) holds a variety of traits, which includes neuropsychiatric disorders ADHD and ASD, neurological disorders, substance use disorders, psychotic disorders (SCZ, BIP), OCD-related disorders (OCD, TS, anorexia nervosa), and the insula measures.

**Figure 6.**
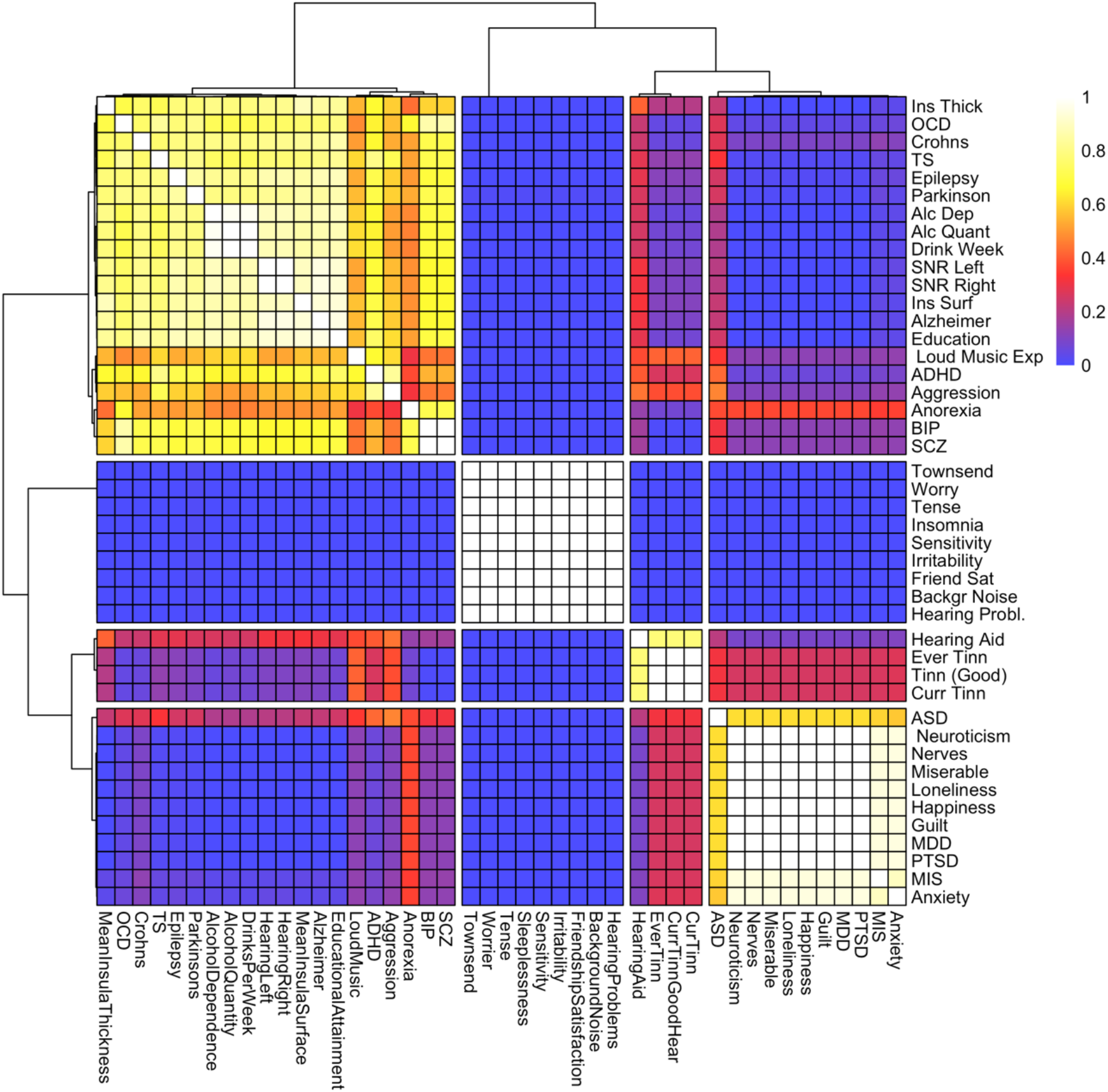
We resampled the genetic correlation matrix using the Monte-Carlo method described in the supplementary methods. For each of 1000 resamplings, we performed the Louvain clustering algorithm. For a majority (69%) of resamplings, this resulted in four clusters. Using all four-cluster solutions, we counted how many times pairs of traits were clustered together, and plotted these numbers on a scale of (0,1). This concordance matrix is plotted above with a reordering of traits according to hierarchical clustering and a grouping of traits into four clusters, which was used to classify the traits as in the graph in figure 6 (main text). The results show that the clustering was highly consistent for many traits across genetic correlation matrix resamples.

## Discussion

Our main aim was to investigate the nature of misophonia as a disorder by functionally annotating a GWAS for misophonia to reveal neurobiological function of risk SNPs, and to calculate and cluster genetic correlations between misophonia and a wide range of other disorders and behavioural traits. As the GWAS for misophonia does not yet exist, we used a proxy in the form of a GWAS of a misophonia symptom assessed in a general population-based sample (23andMe). Two independent locations showed genome-wide significant hits. A highly significant area on chromosome 5 was found intronic to the TENM2 gene (build 38) as well as SNPs in flanking intergenic regions, The second region on chromosome 1 held expression quantitative trait loci (eQTLs) for neural growth factor 1 (NEGR1), a gene strongly related to various variables related to cognition and socioeconomic status, including cognitive performance & educational attainment (Lee et al., 2018), BMI (Pulit et al., 2019), substance and protein intake (Liu et al., 2019; Niarchou et al., 2020), and depression symptoms (Baselmans et al., 2019). TWAS implicated TFB1M, a modulator gene for inherited deafness (Bykhovskaya et al., 2004) and associated with intelligence in GWAS (Lee et al., 2018). These functional annotations did not point consistently to specific risk genes or neural mechanisms, even though the GWAS showed significant SNP-based heritability.

We found that misophonia showed significant genetic correlations with traits and disorders from several categories: misophonia was positively correlated with several assessments of tinnitus (an audiological disorder), with case-control GWAS of PTSD, MDD and generalized anxiety (psychiatric disorders). The strongest correlations were observed with a range of personality traits that broadly fell into two categories, roughly categorized as neuroticism/guilt and irritability/sensitivity trait clusters. In addition, a negative correlation was observed with educational attainment, in line with the functional annotation of the GWAS.

Our findings are largely consistent with the extant literature on comorbid disorders, with some notable deviations. Previous research has reported on comorbidities of misophonia with psychiatric disorders, and on correlations of misophonia with symptoms and personality traits. The reported comorbid were Post Traumatic Stress Disorder (PTSD), Anorexia Nervosa (AN), bulimia nervosa, Attention Deficit/Hyperactivity Disorder (ADHD), and Autism Spectrum Disorder (ASD) (Schröder et al., 2013; Erfanian et al., 2019). The most extensive study of comorbidities to date (Jager et al.; 2020) reported no axis I comorbidity in 72% of patients diagnosed with misophonia and no axis II disorders in 59%, and consisted of mood disorders, Attention Deficit/Hyperactivity Disorder (ADHD), and Autism Spectrum Disorder (ASD). Contrary to the earlier reports (Schröder et al., 2013; Erfanian et al., 2019) no comorbid PTSD was found. For axis II, comorbid obsessive-compulsive personality and borderline personality disorders were observed. Obsessive-compulsive personality *traits*, however, were found in 26% of the patients, and clinical levels of perfectionism were found in over 66% of misophonia sufferers. Jager et al. concluded that misophonia is a distinct nosological entity categorized within obsessive-compulsive spectrum disorders, with symptoms of compulsivity and impulsivity.

The genetic correlation of misophonia with tinnitus is consistent with the disorder first noted in the audiological literature—similar to but different from tinnitus, hyperacusis and phonophobia (Jastreboff and Jastreboff, 2001). As expected (Jager et al.,) Misophonia did not correlate with any of the hearing performance or hearing loss traits, in stark contrast to tinnitus that showed significant correlations with Loud Music Exposure and Hearing Aid. Tinnitus did not correlate strongly with SNR, which may be considered surprising as tinnitus is often associated with hearing loss (Langguth et al., 2013). It is also consistent with findings that people with tinnitus but normal hearing thresholds may have hair cell damage or otherwise affected cochlear regions, suggesting that hearing loss does not necessarily lead to loss of hearing performance that can be detected with clinical tests (Weisz et al., 2006; Job et al., 2007; Langguth et al., 2013). The results may indicate that misophonia is not related to hearing loss, however, methodological issues remain. The triple digit test is not the gold standard, nor the most sensitive measure of hearing sensitivity. More sensitive measures of hearing sensitivity, such as measurement of hearing threshold across the frequency spectrum or measurement of speech-in-noise perception with CVC words, may reveal such a relationship. We note that no GWAS is available of hyperacusis or phonophobia, therefore, our results do not preclude the possibility that other sensory problems than hearing loss play a role in misophonia.

Contrary to the expectations from comorbidity analyses (Jager et al., 2020; Williams et al., 2021), a negative correlation with ASD was observed. The emotional response that defines misophonia may also be found in patients with ASD (Jastreboff and Jastreboff, 2014; Williams et al., 2021), but this is not reflected in an overlap in genomic variation. It has been noted that despite the decreased sound tolerance observed often in ASD, misophonia sufferers with comorbid ASD are a minority of the misophonia cases (3% in Jager et al., 2020), with ASD cases more frequently forming *hyperacusis* (Williams et al., 2021). Nevertheless, over 25% of children with hyperacusis—most of which had clinical ASD—indicated having misophonia symptoms (Amir et al., 2018). Our results suggest that misophonia and ASD are relatively independent disorders with regard to genomic variation, the small protective effect likely being mediated by the positive correlation of ASD with cognition (r_G_(ASD,EA)=0.21). It raises the possibility that two forms of misophonia may exist, one that is mostly driven by conditioning of anger or other negative emotionality to specific trigger sounds moderated by personality traits; the second forming a smaller subgroup that is driven to a greater extent by decreased sound tolerance (Williams et al., 2021), which was not picked up by the current misophonia GWAS in a population-based sample.

Another result that could be considered unexpected is that the positive correlation with aggression was not significant, even though anger and aggressive thoughts are a main symptom of misophonia (Jastreboff and Jastreboff, 2001; Jager et al., 2020). It has been argued, however, that misophonia is based on the feelings of guilt about the evoked irritation and anger rather than behavioural expressions of anger itself that causes the distress (Jager et al., 2020) making the disorder more compulsive than impulsive in character (Eijsker et al., 2020). It should be noted, however, that the GWAS for aggression was relatively small, and future updates of the aggression GWAS may show a significant positive correlation. The GWAS for another impulsive disorder (viz., ADHD) had, however, ample power. The lack of genetic correlation with ADHD—and lack of clustering of misophonia with in the ADHD / aggression cluster—provides further evidence for the disorder not belonging in the impulsive disorders cluster.

Almost no correlation was found with Anorexia (r_G_=0.03, n.s.), in contrast to previous studies into phenotypic comorbidities of misophonia (Erfanian et al., 2019; Kluckow et al., 2014). Lastly, there was barely any correlation with OCD (r_G_ = 0.04, n.s.), even though previous studies did report a link (Wu et al., 2014; Erfanian et al., 2018; Webber et al., 2013). The explanation for this could be found in the distinction between OCD and obsessive compulsive personality disorder (OCPD). For example, Jager et al. (2020) found OCD and OCPD to be comorbid with misophonia, but at a different level: 2.8% of the patients had comorbidity with OCD, while 26% had comorbid symptoms of OCPD. This former prevalence is only slightly higher than population prevalence for OCD (1.6%; den Braber *et al*., 2016), but a substantial increase for OCPD (7.8% in the US; (Grant et al., 2004)). However, to date no GWAS of OCPD has been performed, precluding its use in the current analysis. Nevertheless, the finding that misophonia clusters with psychiatric disorders and related personality dimensions seems to support it either as a highly specific variant of OCPD or a separate personality disorder with strong comorbidity.

Finally, a surprising result was the negative genetic correlation with Educational Attainment, which was significant after correction (r_G_=–0.18, p=3.1 × 10^−7^). Educational Attainment is well-known to correlate with many psychiatric disorders, in part as a non-cognitive indicator of environment or SES (Abdellaoui et al. 2019). The pattern of genetic correlations of misophonia closely mimicked that of MDD, that also showed a small but significant negative correlation with educational attainment. In addition, MDD showed a stronger association with the noncognitive variance in educational attainment than the cognitive variance (Demange *et al*., 2021; figure 4). Again, this resonates well with the interpretation of noncognitive variance in educational attainment as personality related. Another indicator for socioeconomic status— the Townsend Index—did not correlate with misophonia. We therefore do not expect that socioeconomic environmental factors play a substantial role in misophonia symptoms.

The results of the graph clustering concurred with the hierarchical clustering observed in figure 2, placing it in a cluster with MDD, PTSD, Guilt, Nerves, Happiness, Loneliness, Neuroticism, and Anxiety. Monte-Carlo resampling of the genetic correlation matrix showed that this clustering was highly consistent (>95%). This leads us to the conclusion that misophonia may be classified as a psychiatric disorder related to MDD and PTSD, with contributing personality dimensions in the guilt/neuroticism spectrum. In addition, personality dimensions from the irritability cluster contribute to the disorder to a lesser degree. Consistent with the genetic correlation analyses—but inconsistent with clinical observations (Jager et al., 2020)— impulsive disorders/traits like aggression and ADHD do not cluster with misophonia.

The main limitation of the current study is the fact that the GWAS of misophonia was based on a symptom of misophonia rather than a case-control study of misophonia. In addition, the GWAS was limited with a sample size of around 80k subjects and with a limited set of SNPs available (∼600k genotyped SNPs). We were also limited in the availability of published GWAS, which is currently lacking in analyses of psychiatric personality disorders such as OCPD, and of (cognitive) symptoms of inflexibility and perfectionism as part of the part of the OCPD spectrum. As a methodological note, the participation of UK Biobank and 23andMe may come with a participation bias, likely selecting on higher educational attainment. This selection bias may limit the generalizability of the results to the more educated part of the population or may introduce effects in the form of collider bias.

To summarize, our results showed significant effects of several SNPs on a typical misophonia symptom, the hatred for chewing sounds. The TENM2, TMEM256, NEGR1 and TFB1M genes are candidates for mediating the effects. Genetic correlation analysis suggested that misophonia is not merely an audiological disorder related to sensory trauma or hearing loss, but it is related to tinnitus. Misophonia is related to personality traits in the neuroticism cluster, and, to a lesser degree, to the irritability/sensitivity cluster. Finally, misophonia shares genetic etiology with PTSD, MDD and anxiety disorders. Our conclusions may aid DSM classification and could suggest that different therapy approaches are possible for patients classified on the contributing personality dimensions. To further strengthen this conclusion, more research is needed, and the number of GWAS needs to be extended.

## Data Availability

- All data produced in the present study are available upon reasonable request to the authors
- All scripts will be made available upon reasonable request
- All publicly available data downloaded and used in this manuscript and their source links are made available in the supplementary table.
- One non-public dataset was graciously provided by 23andMe, inc. and may be requested by other authors.

## Conflict of interest

The authors declare none.

## Acknowledgements

We would like to thank the research participants and employees of 23andMe for making this work possible. This study was conducted using UK Biobank resources under application number 40310.

## Notes

### Competing Interest Statement

The authors have declared no competing interest.

### Funding Statement

This study did not receive any funding

