## supplemental table 1 for "Genetic evidence for the link of misophonia with psychiatric disorders and personality"

Supplemental table 1: source GWAS used in the analyses

| **Disorder** | **Abbreviation corrplot** | **Abbreviation clusterplot** | **#Cases** | **#Controls** | **Total** | **Observed Prevalence*** | **Observed SNP heritability (SE)** | **References** |
| --- | --- | --- | --- | --- | --- | --- | --- | --- |
| Misophonia | MIS | MIS | 17606 | 63001 | 80607 | 0.280 | 0.1504 (0.0143) | (Fayzullina *et al.*, 2015) |
| Hearing problems with background noise | BackgroundNoise | Back Noise | 125089 | 205670 | 330759 | 0.378 | 0.0826 (0.0039) | http://www.nealelab.is/uk-biobank/ |
| Current Tinnitus with good hearing | CurrTinnGoodHear | Tinn Good |  |  | 93108 |  | 0.0549 (0.0061) | n/a^†^ |
| Current Tinnitus | CurrTinn | Curr Tinn | 45810 | 199181 | 244991 | 0.187 | 0.135 (0.0095) | n/a^†^ |
| Ever Tinnitus | EverTinn | Ever Tinn | 71772 | 173219 | 244991 | 0.293 | 0.1008 (0.0072) | n/a^†^ |
| Hearing Aid User | HearingAid | Hear Aid | 20355 | 369692 | 390047 | 0.052 | 0.0716 (0.0091) | n/a^†^ |
| Hearing Problems | HearingProblems | Hear Probl | 84839 | 239139 | 323978 | 0.262 | 0.0747 (0.0041) | n/a^†^ |
| Hearing test left | HearingLeft | SNR Left | 1251 | 243200 | 244451 |  | 0.4923 (0.0898) | n/a^†^ |
| Hearing test right | HearingRight | SNR Right | 1420 | 243031 | 244451 |  | 0.375 (0.0791) | n/a^†^ |
| Loud Music Exposure | LoudMusic | Loud Mus. | 28762 | 216539 | 245301 | 0.117 | 0.0328 (0.0035) | n/a^†^ |
| Attention Deficit/Hyperactivity Disorder | ADHD | ADHD | 20183 | 35191 | 55374 | 0.028 | 0.1771 (0.0113) | (Demontis *et al.*, 2019) |
| Alcohol Dependence | AlcoholDependence | Alc Dep | 11569 | 34999 | 46568 | 0.248 | 0.1798 (0.0321) | (Walters *et al.*, 2018) |
| Anorexia Nervosa | Anorexia | AN | 16992 | 55525 | 72517 | 0.006 | 0.1261 (0.0083) | (Watson *et al.*, 2019) |
| Anxiety and related disorders | Anxiety | Anx | 7016 | 14745 | 21761 | 0.036 | 0.0754 (0.0285) | (Otowa *et al.*, 2016) |
| Autism Spectrum Disorder | ASD | ASD | 18381 | 27969 | 46350 | 0.007 | 0.1029 (0.0088) | (Grove *et al.*, 2019) |
| Bipolar Disorder | BIP | BIP | 20352 | 31358 | 51710 | 0.010 | 0.2018 (0.0099) | (Stahl *et al.*, 2019) |
| Major Depressive Disorder | MDD | MDD | 127552 | 233763 | 361315 | 0.170 | 0.0846 (0.0035) | Howard et al. (2019) |
| Obsessive Compulsive Disorder | OCD | OCD | 2688 | 7037 | 9725 | 0.015 | 0.2472 (0.0383) | (IOCDF and OCGAS consortia *et al.*, 2018) |
| Post Traumatic Stress Disorder | PTSD | PTSD | 23212 | 151447 | 174659 | 0.011 | 0.0208 (0.0037) | (Nievergelt *et al.*, 2019) |
| Schizophrenia | SCZ | SCZ | 67390 | 67390 | 134780 | 0.010 | 0.1832 (0.0068) | The Schizophrenia Working Group of the Psychiatric Genomics Consortium (2020) |
| Tourette's Syndrome | TS | TS | 4819 | 9488 | 14307 | 0.010 | 0.2227 (0.0255) | Yu et al. (2019) |
| Aggression | Aggression | Aggr |  |  | 18988 |  | 0.0515 (0.0246) | Pappa et al. (2015) |
| Alcohol Quantity | AlcoholQuantity | Alc Quan |  |  | 112117 |  | 0.0798 (0.0061) | Clarke et al. (2017) |
| Drinks per week | DrinksPerWeek | Drink Week |  |  | 941280 |  | 0.0492 (0.0023) | Liu et al. (2019) |
| Friendship Satisfaction | FriendshipSatisfaction | Friend Sat |  |  | 110118 |  | 0.0613 (0.0049) | http://www.nealelab.is/uk-biobank/ |
| Guilty feelings | Guilt | Guilt | 158898 | 415122 | 574020 | 0.277 | 0.0904 (0.0037) | http://www.nealelab.is/uk-biobank/ |
| Happiness | Happiness | Happ. |  |  | 245445 |  | 0.061 (0.0043) | http://www.nealelab.is/uk-biobank/ |
| Irritability | Irritability | Irrit. | 90282 | 232386 | 322668 | 0.280 | 0.1204 (0.0075) | http://www.nealelab.is/uk-biobank/ |
| Loneliness | Loneliness | Loneliness | 81011 | 367945 | 448956 | 0.180 | 0.0383 (0.0021) | http://www.nealelab.is/uk-biobank/ |
| Miserableness | Miserable | Miserable | 235072 | 339563 | 574635 | 0.409 | 0.0992 (0.0043) | http://www.nealelab.is/uk-biobank/ |
| Suffer from 'nerves' | Nerves | Nerves | 112811 | 461822 | 574633 | 0.196 | 0.09 (0.005) | http://www.nealelab.is/uk-biobank/ |
| Neuroticism score | Neuroticism | Neurot. |  |  | 168105 |  | 0.0958 (0.0078) | http://www.nealelab.is/uk-biobank/ |
| Sensitivity/hurt feelings | Sensitivity | Sens. | 182340 | 145492 | 327832 | 0.556 | 0.0981 (0.0051) | http://www.nealelab.is/uk-biobank/ |
| Sleeplessness/insomnia | Insomnia | Insomn. |  |  | 360738 |  | 0.0646 (0.0027) | http://www.nealelab.is/uk-biobank/ |
| Tense/'highly strung' | Tense | Tense | 56012 | 271220 | 327232 | 0.171 | 0.1226 (0.0067) | http://www.nealelab.is/uk-biobank/ |
| Worrier/anxious feelings | Worrier | Worry | 186559 | 142158 | 328717 | 0.568 | 0.1245 (0.0076) | http://www.nealelab.is/uk-biobank/ |
| Alzheimers | Alzheimer | Alzh | 71880 | 383378 | 455258 | 0.055 | 0.0554 (0.0128) | Jansen et al. (2019) |
| Crohn’s Disease | Crohn | Crohn |  |  | 59957 | 0.032 | 0.4491 (0.0412) | De Lange et al. (2017) |
| Educational Attainment | EducationalAttainment | Educ. |  |  | 1131881 |  | 0.0762 (0.0021) | Lee et al., 2018 |
| Epilepsy | Epilepsy | Epilep | 15212 | 29677 | 44889 | 0.007 | 0.4784 (0.046) | The International League Against Epilepsy Consortium on Complex Epilepsies (2018) |
| Mean Insula Surface | MeanInsulaSurface | Ins Surf |  |  | 22824 |  | 0.2921 (0.0286) | Hofer et al. (2019) |
| Mean Insula Thickness | MeanInsulaThickness | Ins Thick |  |  | 22824 |  | 0.1066 (0.0185) | Hofer et al. (2019) |
| Parkinsons | Parkinsons | Park.Dis | 56306 | 1417791 | 1474097 | 0.060 | 0.0184 (0.0019) | Nalls et al. (2019) |
| Townsend deprivation index at recruitment | Townsend | Towns. |  |  | 336798 |  | 0.0355 (0.002) | http://www.nealelab.is/uk-biobank/ |
| Note. ^†^GWAS performed by the authors (see methods). *Prevalence of the disorder/trait not available for ordinal and quantitative scaled variables. | | | | | | |  |  |

Supplemental table 2: Finemapping SNP effects.

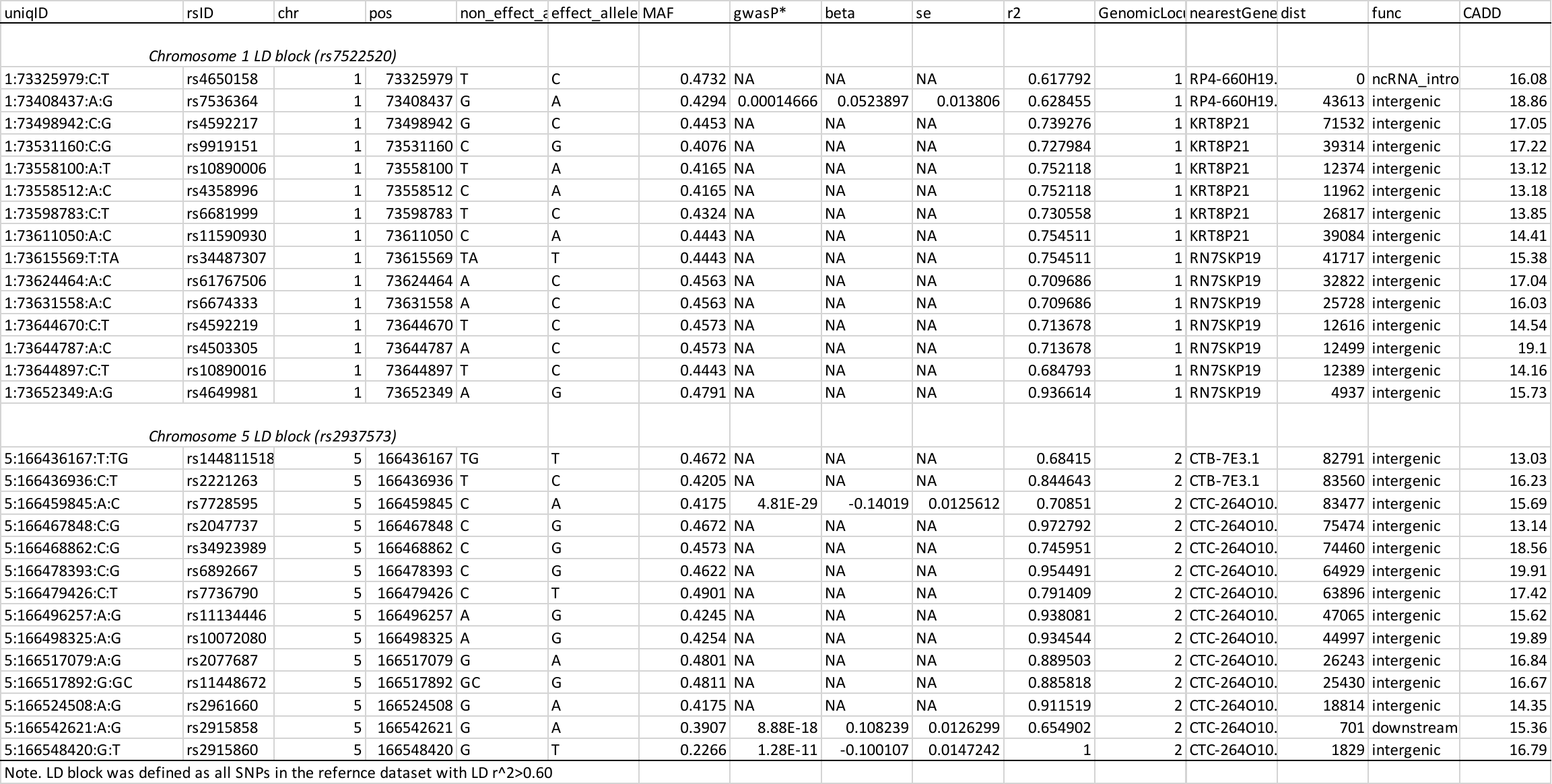
