## Supplementary figures and images for "Genetic evidence for the link of misophonia with psychiatric disorders and personality"

### supplemental table 2

Supplemental table 2: Finemapping SNP effects.

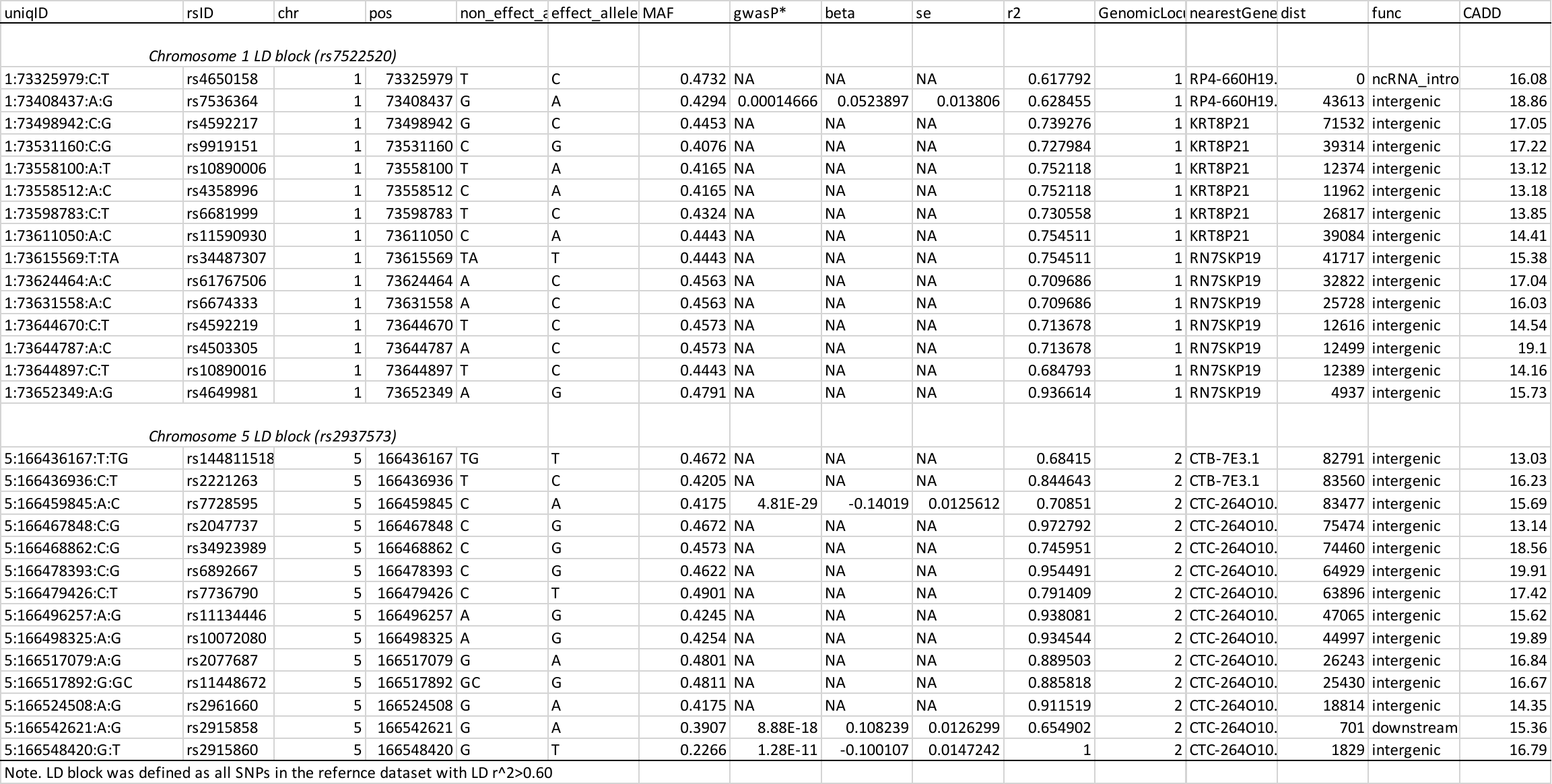
