## supplemental methods for "Genetic evidence for the link of misophonia with psychiatric disorders and personality"

*Monte Carlo sampling approach*

We used LD-score regression implemented in R package Genomic SEM (<https://github.com/GenomicSEM/GenomicSEM>) to obtain the 44x44 r_G_ matrix constructed from the two traits of interest. This r_G_ matrix served as basis for the Genomic SEM models as shown in the main text. The unique elements of the r_G_ matrix (i.e., the SNP-h2 scores from the diagonal and the covariances on the lower off-diagonal elements) were stored in a vector of length (N*(N+1)/2) = 990. In genomic SEM, standard errors of all estimates (factor loadings, residual variances, regression paths and correlations) are based on the variability of the r_G_ matrix. This variability is calculated duding LD score regression with a jacknife method: SNPs are sorted by Chromosome and Basepair positions, split into 200 sets (adjustable figure), and each of these sets is removed from the full set in turn. Then, the r_G_ is recalculated. Each of these 200 r_G_ matrices varies slightly around estimates for the full data.

The result from the jacknife procedure is a (co)variance matrices between all 990 unique h^2^ and r_G_ estimates, i.e. a 990-by-990 covariance matrix. Note that the covariability between estimates tends to be substantial. It is important to include this covariability of the estimates in the sampling, because assuming that r_G_ estimation sampling variability is independent will lead to substantial loss of statistical power and incorrect inference.

Armed with the point estimate vector and the covariance matrix of these estimates, we used R function mvrnorm (MASS package) to create random samples that keeps all dependencies intact. Restructuring each sample back to an r_G_ matrix, we re-estimated the Louvain clustering. We then calculated the proportion of times traits clustered with each other, particularly for the consistency of clustering of traits with misophonia.
